# End-to-End PET/CT Interpretation and Quantification with an LLM-Orchestrated AI Agent: A Real-World Pilot Study

**DOI:** 10.64898/2026.02.21.26346798

**Authors:** Hongyoon Choi, Sungwoo Bae, Kwon Joong Na

## Abstract

**Background:** Although deep learning models have improved individual PET analysis, image processing and quantification tasks, end-to-end automation from raw DICOM to quantitative clinical reporting remains limited, particularly in heterogeneous real-world settings.

**Methods:** As a proof-of-concept, an autonomous large language model (LLM)-orchestrated multi-tool agent for end-to-end PET/CT interpretation was developed. A reasoning-based text LLM selected appropriate series from raw DICOM, coordinated registration and SUV conversion, invoked segmentation and detection tools, generated maximum-intensity projections, called a vision-enabled LLM for interpretation, and synthesized structured draft reports. The system was retrospectively evaluated in 170 patients undergoing baseline FDG PET/CT for lung cancer staging, using expert reports as reference.

**Results:** The agent successfully completed the full end-to-end workflow from raw DICOM selection to structured draft report generation without human intervention in all 170 examinations. Primary tumor detection achieved 100% sensitivity. For nodal involvement, sensitivity was 84.8% and specificity was 39.4%, whereas distant metastasis detection showed 70.2% sensitivity and 65.0% specificity. Discrepancy analysis of 58 nodal and 57 metastatic mismatch cases revealed systematic false-positive findings related to reactive or physiologic uptake and false-negative findings involving small-volume or anatomically atypical metastases.

**Conclusion:** LLM-orchestrated PET/CT agents can enable workflow-level automation from raw DICOM to quantification and structured draft reporting under real-world conditions. Although primary tumor detection was highly reliable, nodal and metastatic assessment revealed systematic limitations, supporting a collaborative role with continued expert oversight in complex clinical scenarios.

## INTRODUCTION

PET/CT provides both qualitative and quantitative information across a wide range of clinical applications, including standardized uptake values (SUVs) and tumor burden metrics [1]. Recent advances in task-specific artificial intelligence (AI) have enabled automated components of PET analysis [2,3], such as organ segmentation for regional quantification and lesion-focused measurements including metabolic tumor volume [4]. However, real-world clinical PET/CT interpretation requires a tightly coupled, multi-step workflow that extends beyond isolated segmentation or quantification tasks [5]. Routine clinical interpretation involves a quantitative imaging process that requires multiple image-processing steps, which may vary across centers and clinical scenarios. These steps include selecting appropriate image series from raw DICOM studies, performing registration and resampling, and converting raw counts to SUVs using patient and injection metadata. Following this quantitative preprocessing, nuclear medicine physicians identify and segment relevant lesions—either semi-automatically or manually—assess whole-body disease burden, and synthesize structured reports tailored to the specific clinical question [5]. Despite recent progress in deep learning–based tools, these workflow-level steps remain largely performed or supervised by expert readers [2,3].

This gap becomes particularly pronounced in real-world clinical environments, where PET/CT data are highly heterogeneous across scanners, acquisition protocols, reconstruction settings, SUV scaling, and metadata completeness. Such variability poses a major barrier to fully automated interpretation, as an effective system must robustly select appropriate series, resolve inconsistencies in DICOM information, and coordinate multiple downstream tools under uncertain conditions. However, most existing segmentation and detection approaches are developed on curated datasets with ‘structured’ situations and do not address these workflow-level challenges [6-9]. Large language and multimodal models may help bridge this gap by reasoning over text, images, and metadata to orchestrate complex imaging pipelines [10,11], although their clinical deployment requires careful definition of appropriate roles and oversight [12,13].

In this study, we suggested and evaluated an autonomous, large language model (LLM)– orchestrated multi-tool agent designed to automate end-to-end FDG PET/CT interpretation as a proof-of-concept study. Unlike conventional single-task models, the proposed system functions as a cognitive orchestrator that coordinates image processing, quantitative analysis, lesion detection, and structured report generation in a manner aligned with real-world clinical workflows [14,15]. Guided by simple text prompts, the agent dynamically selects appropriate PET/CT series, invokes specialized tools, and generates quantitative outputs and draft reports interactively [14]. We retrospectively validated the system in a cohort of 170 patients undergoing baseline lung cancer staging, assessing workflow-level robustness, diagnostic performance, and the clinical acceptability of generated reports. Our findings illustrate the potential role of general-purpose AI agents as workflow-level assistants that provide structured analytical baselines while maintaining the need for expert oversight in complex clinical scenarios [13,15].

## MATERIALS AND METHODS

### Study design

This retrospective single-center study utilized clinical PET/CT imaging data obtained from the institutional Clinical Data Warehouse. We identified patients who underwent routine FDG PET/CT for lung cancer staging in 2018 and extracted a cohort of 170 cases for pilot evaluation of the proposed workflow-based approach. In addition, publicly available PET datasets from The Cancer Imaging Archive (TCIA; https://www.cancerimagingarchive.net/) [16] were used for application tests. Institutional Review Board approval was obtained (IRB No. 2105-157-1221), and the requirement for written informed consent was waived because of the retrospective design and the use of deidentified data.

### FDG PET dataset

The study cohort consisted of 170 unique adult patients who underwent baseline FDG PET/CT for the diagnostic work-up or initial staging of suspected or newly diagnosed primary lung cancer. Baseline staging scans were defined as the first PET/CT examination performed prior to any systemic or local oncologic treatment. Studies performed for restaging, treatment response assessment, or surveillance were excluded. Additional exclusion criteria included incomplete PET or CT series, and corrupted DICOM files that could not be parsed. All examinations were acquired under routine clinical protocols on heterogeneous PET/CT systems. Variability in series naming conventions, reconstruction parameters, voxel sizes, and SUV scaling reflected real-world clinical conditions.

For the single-center cohort, patients fasted for at least 8 h, and blood glucose levels were measured before tracer injection. [^18^F]FDG was administered at a dose of 5.18 MBq/kg when blood glucose was below 200 mg/dL. After voiding, PET/CT imaging was performed approximately 60 min after injection. Scans were acquired using 3 dedicated PET/CT systems (Biograph TruePoint 40, Biograph mCT 40, and Biograph mCT 64; Siemens Healthineers, Erlangen, Germany). Low-dose non–contrast-enhanced CT was obtained from the skull base to the proximal thigh (120 kVp; matrix size, 512 × 512; slice thickness, 5 mm for TruePoint 40 and 3 mm for mCT systems). Subsequently, PET emission imaging was performed, with acquisition times of 2 min/bed (TruePoint) and 1 min/bed (mCT systems).

### LLM-orchestrated agent architecture

The proposed system was designed using a three-layer architecture consisting of (1) a cognitive control layer, (2) a tool abstraction layer, and (3) an execution layer [10,14]. The overall architecture and data flow from raw DICOM input to quantitative report generation are illustrated in **Figure 1**. The cognitive control layer was implemented using a text-based LLM (Gemini-3-flash-preview) [17], which functioned as the workflow orchestrator. A predefined system prompt specified its role as a PET/CT interpretation assistant responsible for planning the analysis, selecting appropriate tools, monitoring intermediate outputs, and generating structured draft reports. Task-specific prompt templates guided key steps, including DICOM series selection, image preprocessing, lesion quantification, and SUV metric computation. Tool interactions were performed through a structured schema defining tool names, required inputs, and expected outputs. The orchestrator employed an iterative reasoning framework in which each tool call followed a Thought–Action– Observation sequence, enabling stepwise planning and validation [18]. All reasoning steps and tool invocations were logged. The tool abstraction layer translated high-level requests from the orchestrator (e.g., lesion segmentation or background SUV calculation) into executable functions. Tool metadata defined input modalities (PET, CT, masks, or derived images), file formats, and expected behavior, allowing the system to detect failures or empty outputs and adapt accordingly. The execution layer consisted of Python-based modules responsible for DICOM handling, image registration and resampling, SUV computation, visualization, and deep learning–based segmentation and detection.

**Figure 1.**
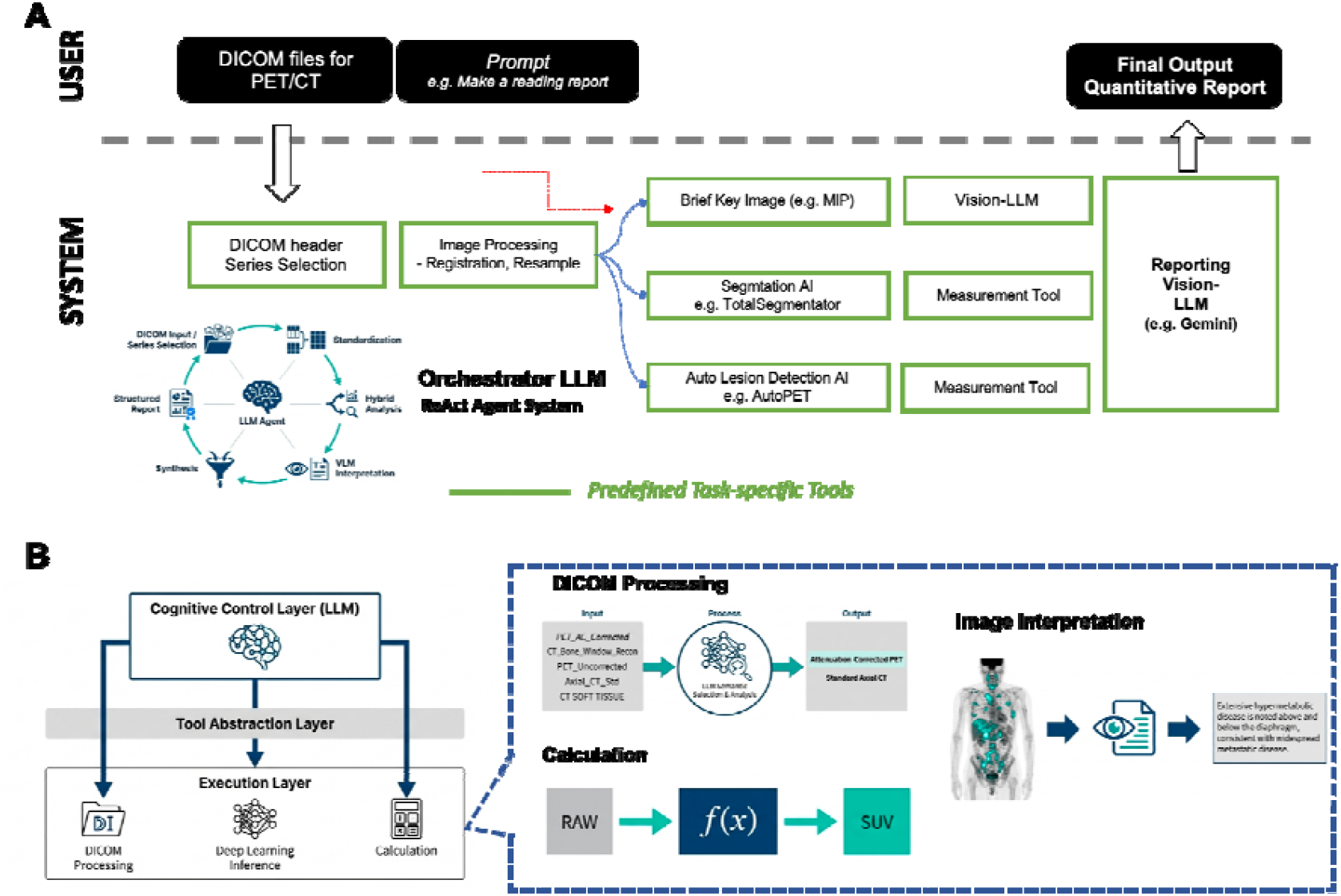
Architecture of the LLM-orchestrated multi-tool PET/CT workflow. Schematic overview of the proposed LLM-orchestrated agent architecture for end-to-end PET/CT interpretation and quantitative reporting. (A) Workflow from raw DICOM input to final quantitative report generation. Clinical PET/CT DICOM files are provided together with a user prompt. The cognitive control layer (LLM) performs DICOM header parsing and series selection, followed by image preprocessing including registration and resampling by invoking tools. Based on the task, the orchestrator dynamically invokes predefined task-specific tools, including key image generation (e.g., maximum-intensity projection), organ segmentation (e.g., TotalSegmentator), automated lesion detection (e.g., AutoPET), and quantitative measurement modules. Outputs from these tools are integrated and forwarded to a reporting Vision-LLM, which generates a structured narrative and quantitative report. (B) Three-layer system design comprising (1) a cognitive control layer responsible for planning, tool selection, and validation; (2) a tool abstraction layer that maps high-level commands to executable modules; and (3) an execution layer that performs DICOM handling, deep learning inference, and quantitative calculations.

### Imaging Tools, Quantification, and Workflow Execution

The agent integrated multiple task-specific imaging tools to enable end-to-end PET/CT interpretation and quantification. The primary aim of this study was not to develop new models for individual tasks such as lesion segmentation, organ delineation, or quantitative measurement. Rather, the goal was to evaluate a workflow-level AI agent capable of interactively orchestrating existing specialized tools in response to user prompts, thereby more closely resembling routine clinical PET/CT interpretation processes.

Whole-body PET lesion segmentation was performed using an AutoPET nnU-Net–based 3D model trained for volumetric tumor delineation [7-10] (https://github.com/MIC-DKFZ/autopet-3-submission). Resulting masks were used to compute quantitative metrics including SUVmax, metabolic tumor volume (MTV), and total lesion glycolysis (TLG) [4]. Organ segmentation was performed on CT using TotalSegmentator to obtain major anatomic structures and reference regions [19].

A vision-enabled LLM (Vision-LLM; Gemini-3-flash-preview) was used for image-based interpretation [10,17]. The orchestrator generated standardized key visualizations, including whole-body PET maximum-intensity projections, multiplanar PET/CT fusion slices, and lesion or organ mask overlays, and prompted the Vision-LLM to provide lesion-level summaries of location, uptake intensity, and suspected category (primary, nodal, metastatic, or physiologic). These outputs were incorporated into staging assessment and structured report generation.

For each study, the orchestrator parsed raw DICOM series and selected an attenuation-corrected whole-body PET series with the corresponding CT. PET and CT volumes were aligned and resampled into a common space to ensure consistent fusion and quantification. SUV values were computed voxel-wise using injected activity, patient weight, and acquisition timing extracted from DICOM metadata [1]. Lesion- and organ-level quantitative summaries were then automatically generated and synthesized into staging-oriented draft reports [5].

Tool calls were monitored using predefined error-detection rules, including checks for missing outputs, invalid quantitative ranges, or runtime failures. When lesion segmentation failed or yielded empty masks, the system applied fallback strategies, such as relying on Vision-LLM interpretation of maximum-intensity projections and fusion images for staging decision. All intermediate reasoning steps, tool inputs and outputs, and error events were logged. A subset of cases was selected for qualitative analysis of the agent’s workflow traces.

### Mapping to staging labels and reference standard

For each patient, the original clinical PET/CT report served as the reference standard. Reports were reviewed to extract patient-level labels for primary tumor presence, nodal involvement (N+ vs N0), and distant metastasis (M1 vs M0). Agent outputs were mapped to the same categories using predefined operational criteria. Primary tumor presence was defined as at least one FDG-avid lesion within the lung parenchyma or hilum classified as primary. Nodal involvement was defined as any lesion categorized as nodal disease in mediastinal, hilar, supraclavicular, or other nodal stations. Distant metastasis was defined as FDG-avid lesions outside the thoracic nodal basin labeled as metastatic, including involvement of bone, liver, adrenal glands, brain, or other organs. Discrepancies between agent-derived and reference labels were recorded and subsequently categorized as false-positive or false-negative patterns for nodal and metastatic disease.

### Evaluation and statistical analysis

Workflow-level feasibility was defined as the proportion of examinations in which the agent completed the full end-to-end pipeline from DICOM input to draft report generation without human intervention. Patient-level diagnostic performance for primary tumor detection, nodal involvement, and distant metastasis was evaluated using sensitivity, specificity, accuracy, positive predictive value, and negative predictive value, with the clinical report serving as the reference standard. Confusion matrices were generated for each task.

## RESULTS

### Reactive orchestrator-based outputs

The agent was tested across multiple clinically relevant prompt-driven scenarios, including (1) qualitative draft reporting, (2) organ-based SUV measurement, and (3) quantitative lesion reporting. For qualitative interpretation, the prompt “Interpret the PET scan and draft a report for any abnormalities” triggered the agent to generate whole-body maximum-intensity projections (MIPs) and request Vision-LLM–based assessment of abnormal FDG uptake patterns (**Figure 2A**).

**Figure 2.**
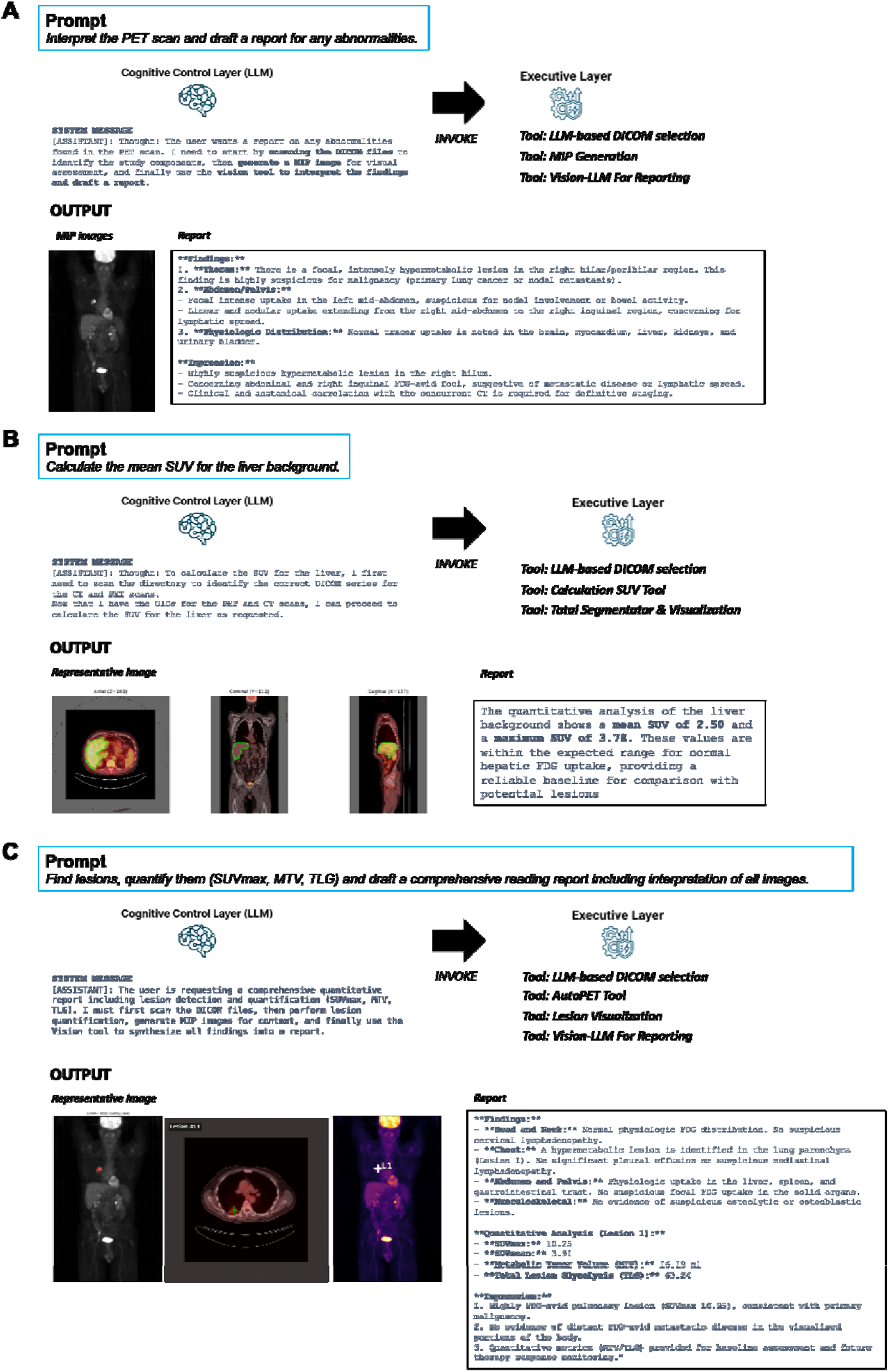
Prompt-driven LLM-agent orchestration for qualitative interpretation and quantitative PET/CT analysis. This figure illustrates representative prompt-driven workflows of the LLM-orchestrated PET/CT agent under three clinically relevant scenarios. (A) Qualitative draft reporting. When prompted to “Interpret the PET scan and draft a report for any abnormalities,” the Cognitive Control Layer (LLM) first selects the appropriate PET/CT DICOM series and generates a whole-body maximum-intensity projection (MIP). The orchestrator then invokes a Vision-LLM to interpret abnormal FDG uptake and produce a structured narrative report. The output includes representative MIP images and a text report summarizing suspected primary and secondary lesions and overall disease impression. (B) Organ-based quantitative SUV computation. In response to the prompt “Calculate the mean SUV for the liver background,” the orchestrator selects the relevant PET/CT series, applies CT-based organ segmentation (TotalSegmentator), and computes SUV metrics from the segmented hepatic parenchyma. Representative axial, coronal, and sagittal PET/CT fusion images with liver masks are shown, along with the automatically generated quantitative summary (mean SUV and maximum SUV) and narrative interpretation. (C) End-to-end lesion detection, quantification, and structured reporting. For a comprehensive prompt requesting lesion detection and quantitative analysis (SUVmax, metabolic tumor volume [MTV], total lesion glycolysis [TLG]), the orchestrator coordinates AutoPET-based whole-body lesion segmentation, lesion visualization, quantitative metric computation, and Vision-LLM–assisted reporting. The output includes lesion overlays on PET images and a structured report summarizing regional findings, per-lesion quantitative indices, and overall staging-oriented interpretation.

**Figure 3.**
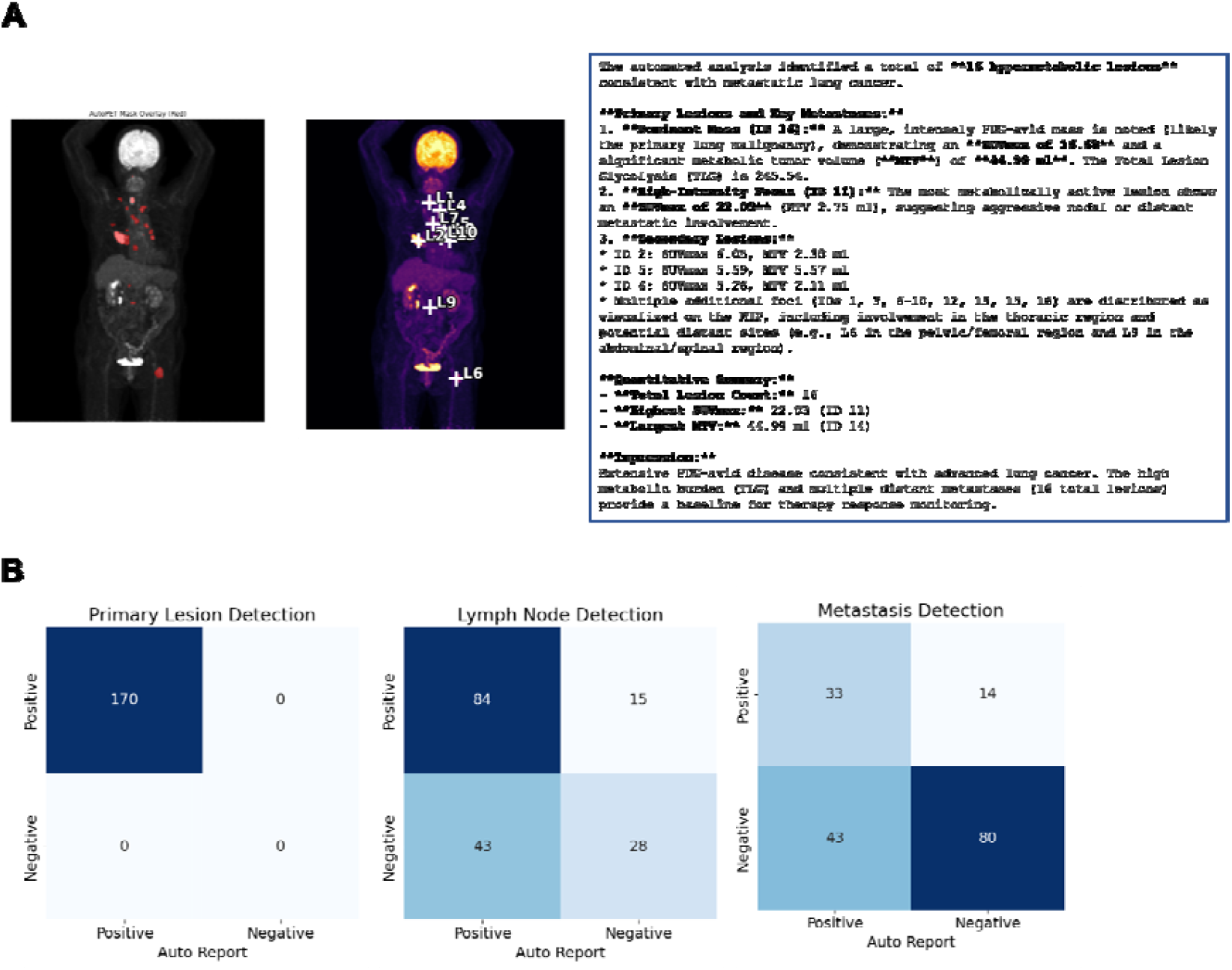
Representative lesion detection outputs and patient-level staging performance. (A) Examples of automated whole-body lesion detection and visualization. AutoPET-based segmentation masks (red overlays) projected on whole-body PET images, demonstrating detected FDG-avid lesions. Representative whole-body PET maximum-intensity projection (MIP) with automatically labeled lesion indices (L1–L10), illustrating multi-lesion identification and spatial distribution. (B) Patient-level confusion matrices comparing agent-generated staging outputs with expert-interpreted clinical PET/CT reports for (left) primary tumor detection, (middle) nodal involvement (N+ vs N0), and (right) distant metastasis (M1 vs M0). For primary tumor detection, the agent achieved 100% sensitivity (170/170). For nodal staging, sensitivity was 84.8% (84/99) and specificity was 39.4% (28/71). For distant metastasis detection, sensitivity was 70.2% (33/47) and specificity was 65.0% (80/123).

For quantitative tasks, the agent dynamically invoked task-specific imaging tools, including TotalSegmentator for organ delineation [19] (**Figure 2B**) and an AutoPET lesion segmentation model for tumor quantification according to the prompts [6-8]. Using these outputs, the system automatically computed standard PET metrics such as lesion SUV and MTV (**Figure 2C**). An example of the agent’s interactive execution, including its reasoning process and tool invocation guided by system prompts, is provided in **Supplementary Video 1**.

End-to-end execution of the predefined workflow —from DICOM loading and series selection through registration/resampling, SUV conversion, segmentation, Vision-LLM consultation, and structured draft report generation—was successfully completed in the majority of examinations. Failures were rare (one case among 170 cases of the cohort) and were primarily attributable to DICOM or metadata inconsistencies, such as insufficient information for SUV calculation. In cases where quantitative lesion tools failed, the agent reverted to a qualitative interpretation mode using MIP-based Vision-LLM assessment to generate clinically meaningful draft reports (**Supplementary Figure 1**).

### Patient-Level Staging Performance

Using expert-interpreted real clinical PET/CT reports as the reference standard, the agent’s staging outputs were evaluated at the patient level for primary tumor detection, nodal involvement, and distant metastasis. An example case is presented in **Figure 4A**, illustrating lesion detection and interpretation through an end-to-end pipeline, from raw DICOM images to the generated narrative report. Confusion matrices and summary performance metrics for these staging tasks are provided in **Figure 4B**.

Patient-level staging performance was assessed against expert clinical PET/CT reports (**Figure 4**). For primary tumor detection, the agent identified all primary lung cancers correctly, achieving a sensitivity of 100% (170/170). For nodal staging, the agent demonstrated high sensitivity for clinically significant lymph node involvement (84.8%, 84/99) but limited specificity (39.4%, 28/71), reflecting frequent false-positive nodal calls. Discrepancy analysis showed that most false positives arose from low-grade FDG uptake in hilar or mediastinal lymph nodes that were interpreted as reactive or physiologic in ‘ground-truth’ expert reports. False-negative nodal cases were less common and typically involved small or mildly avid interlobar, paratracheal, or supraclavicular nodes that were omitted from the automated report. For distant metastasis detection, the agent achieved moderate sensitivity (70.2%, 33/47) and specificity (65.0%, 80/123), with an overall accuracy of 66.5%. Among the metastatic mismatches, false-positive M1 classifications were frequently driven by physiologic uptake in the bowel or pelvis and benign osseous uptake related to degenerative or traumatic changes. In contrast, false negatives most often involved small-volume metastases at anatomically challenging or atypical sites, including subtle bone lesions, pleural seeding, adrenal involvement, thyroid nodules, or intracranial metastases. All false-positive and false-negative cases identified in the discrepancy analysis are summarized in **Supplementary Table 1**.

Together, these discrepancy patterns indicate that the agent provides clinically meaningful staging support but remains vulnerable to borderline nodal uptake and physiologic mimics, underscoring the need for expert oversight in high-risk scenarios.

## DISCUSSION

This study demonstrates the feasibility of an LLM-orchestrated multi-tool agent capable of executing an end-to-end FDG PET/CT staging workflow directly from heterogeneous clinical DICOM data. Unlike prior applications of large language models in nuclear medicine, which primarily operate on pre-existing text reports or structured outputs [20,21], the present system was designed to coordinate image processing, quantitative computation, segmentation tools, and vision-based interpretation within a unified workflow [22]. As a real-world application scenario, in a cohort of 170 baseline lung cancer staging examinations, the agent completed the full pipeline, indicating that workflow-level automation is technically achievable under real-world imaging variability [14,15].

A key contribution of this work lies in shifting the focus from isolated task-specific performance to orchestration across the entire PET/CT interpretation process. Clinical PET reading requires coordinated execution of series selection, registration, SUV conversion, lesion and organ segmentation, and structured reporting. The cognitive control layer by LLM enabled dynamic tool selection and validation, including fallback strategies when segmentation or metadata-dependent SUV computation failed. This layered architecture mirrors how human readers adapt to incomplete metadata or inconsistent image series and supports the concept of LLMs functioning as supervisory “control layers” rather than standalone diagnostic models [10,14].

Diagnostic performance varied across staging components and was influenced in part by the capabilities of the final Vision-LLM responsible for image interpretation and report generation. Importantly, the objective of this AI agent system was not solely to optimize staging accuracy, but to evaluate feasibility in real-world, end-to-end clinical workflows that emulate the interactive reasoning and quantitative processes of human PET interpretation. Nevertheless, primary lung tumor detection achieved 100% sensitivity in this cohort, reflecting the robustness of dominant-lesion identification when combining segmentation outputs and vision-based interpretation [23]. In contrast, nodal and distant metastatic classification demonstrated moderate performance with asymmetric error profiles. Nodal staging was characterized by relatively high sensitivity but low specificity, driven largely by overcalling of low-grade FDG uptake in hilar or mediastinal lymph nodes that were interpreted as reactive or physiologic in expert reports. For distant metastasis, false-positive findings most frequently involved physiologic bowel or pelvic uptake and benign osseous changes, whereas false negatives clustered in small-volume or anatomically atypical metastases, including subtle bone lesions, pleural seeding, adrenal involvement, thyroid nodules, and intracranial disease. These discrepancy patterns were systematic rather than random and reflect known limitations of PET/CT segmentation and interpretation in heterogeneous or borderline uptake scenarios [6,8,24]. In this regard, the goal of the current AI agent is not to replace human expertise in PET/CT interpretation. Rather, such systems should be viewed as workflow-level assistants that can support physicians and researchers by automating repetitive preprocessing and quantitative tasks, providing preliminary lesion measurements, and enabling scalable analysis across large and heterogeneous imaging cohorts.

Importantly, the agent’s behavior illustrates that orchestration does not eliminate the intrinsic strengths and weaknesses of underlying task-specific models. Instead, the LLM layer integrates their outputs and can mitigate some failures through reasoning and fallback strategies, but it cannot fully compensate for domain-generalization limitations or ambiguous uptake patterns [6-8]. The system therefore functions most appropriately as a workflow assistant rather than an autonomous reader. The observed qualitative report acceptability further supports this positioning, with stronger performance in high-burden or straightforward cases and lower reliability in borderline nodal or metastatic scenarios.

One important point is that the primary objective of this study was not to develop or benchmark the best-performing vision-enabled LLM for PET/CT interpretation. Rather, the main contribution was to demonstrate the feasibility of an end-to-end workflow-level agent that can autonomously coordinate quantitative and qualitative PET/CT interpretation steps under heterogeneous real-world clinical conditions. In this framework, the interpretive performance of the system is partly dependent on the underlying Vision-LLM component, and future integration with emerging specialized medical vision-language foundation models may further improve diagnostic robustness and lesion characterization (e.g., Med-Gemma, HuLu-Med) [25,26]. Notably, such improvements could be achieved by substituting or upgrading the Vision-LLM module within the same orchestration architecture, allowing the agent to remain interactive and adaptable across diverse clinical scenarios and quantification purposes.

Methodologically, this study extends AI agent concepts beyond text-only augmentation to direct coordination of image-based tools and quantitative pipelines. By integrating autoPET lesion segmentation and CT-based organ masks within a unified orchestration framework [8,9,19], the system demonstrates how established algorithms can be assembled into a clinically oriented, quantification-ready workflow without retraining each component. This approach may provide a scalable pathway for incorporating multiple validated tools into routine clinical environments while preserving expert oversight [13,15].

Several limitations must be acknowledged. The study is retrospective, and quantitative analysis was restricted to baseline FDG PET/CT for cancer staging. The reference standard in this study was derived from routine clinical PET/CT reports, reflecting the practical goal of evaluating the agent’s performance within an end-to-end real-world interpretation framework. System performance remains dependent on the domain generalizability of the underlying segmentation models and on the completeness and consistency of DICOM metadata, which continues to represent a major barrier to fully automated clinical pipelines [6,8]. In addition, the clinical impact on reading time, decision-making, or patient outcomes was not evaluated. Future work should validate workflow-level agents across multiple institutions, tracers, and disease indications, and should explore integration with next-generation multimodal models capable of deeper spatial reasoning [27,28]]. Standardized benchmarks for end-to-end imaging agents, complementing existing segmentation challenges [6,8], will be necessary to rigorously assess robustness under heterogeneous clinical conditions.

In conclusion, an LLM-orchestrated multi-tool agent can automate most components of the FDG PET/CT staging workflow in real-world heterogeneous data. While primary tumor detection approached expert-level sensitivity, nodal and metastatic assessment revealed characteristic, interpretable error patterns. These findings support the role of general-purpose AI agents as structured workflow assistants that enhance quantitative consistency and draft reporting while maintaining essential expert oversight in complex clinical scenarios.

## DISCLOSURE

### Data availability

Due to personal information protection policies, the complete datasets of reading reports are not available outside the hospital server. Sample data are included in the supplementary materials and their related contents can be provided by the corresponding author upon reasonable request.

### Ethics approval

The retrospective analysis of human data and the waiver of informed consent were approved by the Institutional Review Board of the Seoul National University Hospital.

### Consent to publish

Not applicable.

## REFERENCES

1. Boellaard R, Delgado-Bolton R, Oyen WJG, et al. FDG PET/CT: EANM procedure guidelines for tumour imaging: version 2.0. Eur J Nucl Med Mol Imaging. 2015;42:328–354.

2. Litjens G, Kooi T, Bejnordi BE, et al. A survey on deep learning in medical image analysis. Med Image Anal. 2017;42:60–88.

3. Visvikis D, Cheze Le Rest C, Jaouen V, Hatt M. Artificial intelligence, machine (deep) learning and radio(geno)mics: definitions and nuclear medicine imaging applications. Eur J Nucl Med Mol Imaging. 2019;46:2630–2637.

4. Larson SM, Erdi Y, Akhurst T, et al. Tumor treatment response based on visual and quantitative changes in global tumor glycolysis using PET-FDG imaging. Clin Positron Imaging. 1999;2:159–171.

5. Niederkohr RD, Greenspan BS, Prior JO, et al. Reporting guidance for oncologic 18F-FDG PET/CT imaging. J Nucl Med. 2013;54:756–761.

6. Gatidis S, Hepp T, Früh M, et al. Results from the autoPET challenge on fully automated lesion segmentation in oncologic PET/CT imaging. Nat Mach Intell. 2024;6:1396–1405.

7. Rokuss M, Kovacs B, Kirchhoff Y, et al. From FDG to PSMA: a hitchhiker’s guide to multitracer, multicenter lesion segmentation in PET/CT imaging. arXiv preprint. 2024;arXiv:2409.09478.

8. Dexl J, Gatidis S, Früh M, et al. AutoPET challenge on fully automated lesion segmentation in oncologic PET/CT imaging, part 2: domain generalization. J Nucl Med. 2025. doi:10.2967/jnumed.125.270260.

9. Isensee F, Jaeger PF, Kohl SAA, Petersen J, Maier-Hein KH. nnU-Net: a self-configuring method for deep learning-based biomedical image segmentation. Nat Methods. 2021;18:203–211.

10. Bradshaw TJ, Tie X, Warner J, et al. Large language models and large multimodal models in medical imaging: a primer for physicians. J Nucl Med. 2025;66:173–182.

11. Singhal K, Azizi S, Tu T, et al. Large language models encode clinical knowledge. Nature. 2023;620:172–180.

12. Hosny A, Parmar C, Quackenbush J, Schwartz LH, Aerts HJWL. Artificial intelligence in radiology. Nat Rev Cancer. 2018;18:500–510.

13. Topol EJ. High-performance medicine: the convergence of human and artificial intelligence. Nat Med. 2019;25:44–56.

14. Koçak B, Meşe İ. AI agents in radiology: toward autonomous and adaptive intelligence. Diagn Interv Radiol. 2025. doi:10.4274/dir.2025.253470.

15. Freyer O, Jayabalan S, Kather JN, et al. Overcoming regulatory barriers to the implementation of AI agents in healthcare. Nat Med. 2025;31:3239–3243.

16. Clark K, Vendt B, Smith K, et al. The Cancer Imaging Archive (TCIA): maintaining and operating a public information repository. J Digit Imaging. 2013;26:1045–1057.

17. Saab K, Tu T, Weng WH, et al. Capabilities of Gemini models in medicine. arXiv preprint. 2024;arXiv:2404.18416.

18. Yao S, Zhao J, Yu D, et al. ReAct: synergizing reasoning and acting in language models. Paper presented at: ICLR 2023; Kigali, Rwanda.

19. Wasserthal J, Breit HC, Meyer MT, et al. TotalSegmentator: robust segmentation of 104 anatomic structures in CT images. Radiol Artif Intell. 2023;5:e230024.

20. Choi H, Lee D, Kang YK, Suh M. Empowering PET imaging reporting with retrieval-augmented large language models and reading reports database: a pilot single center study. Eur J Nucl Med Mol Imaging. 2025;52:2452–2462.

21. Al Zaabi A, Alshibli R, AlAmri A, et al. Trends and trajectories in the rise of large language models in radiology: scoping review. JMIR Med Inform. 2025;13:e78041.

22. Teo ZL, Thirunavukarasu AJ, Elangovan K, et al. Generative artificial intelligence in medicine. Nat Med. 2025;31:36–44.

23. Sibille L, Seifert R, Avramovic N, et al. 18F-FDG PET/CT uptake classification in lymphoma and lung cancer by using deep learning with convolutional neural networks. J Nucl Med. 2020;61:46–52.

24. Kandathil A, Kay FU, Butt YM, Wachsmann JW, Subramaniam RM. Role of FDG PET/CT in the eighth edition of TNM staging of non-small cell lung cancer. RadioGraphics. 2018;38:2134–2149.

25. Sellergren AB, Tanno R, Thakoor K, et al. MedGemma: an open medical foundation model collection for multimodal AI. arXiv preprint. 2025. arXiv:2507.05201.

26. Jiang S, Wang Y, Li Z, et al. Hulu-Med: a transparent generalist model towards holistic medical vision-language understanding. arXiv preprint. 2025. arXiv:2510.08668.

27. Li M, Lin F, Hu Y, et al. Towards a holistic framework for multimodal large language models in three-dimensional brain CT report generation. Nat Commun. 2025;16:2258.

28. Ryu JS, Kang H, Chu Y, Yang S. Vision-language foundation models for medical imaging: a review of current practices and innovations. Biomed Eng Lett. 2025;15:809–830.

