## Supplementary Figure1 for "End-to-End PET/CT Interpretation and Quantification with an LLM-Orchestrated AI Agent: A Real-World Pilot Study"

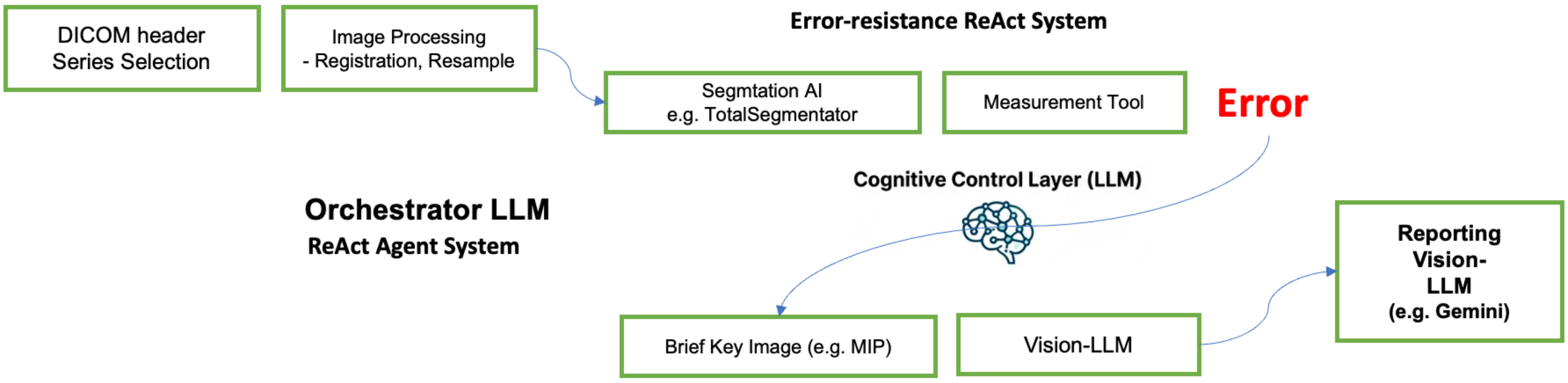


```
[ASSISTANT]: Thought: DICOM files have been identified. Now I need to quantify the lesions using the AutoPET-inspired tool to get SUV
metrics and lesion masks.
Attempting AutoPET-3 Inference...
Preparing explicitly aligned 4.0mm Isotropic data (Union Canvas) to optimize for GPU memory...
AutoPET failed: Could not decompose affine:
[…]
[ASSISTANT]: Thought: The QUANTIFY_LESION tool failed due to a geometric decomposition error in the DICOM headers. To proceed with the
"Full Quantitative Report," I will calculate the SUV for the liver to provide a reference background (Deauville-like context) and
generate MIP images so I can visually interpret the scan and provide the requested report.
Observation: Error SUV: Could not decompose affine:
[…]
[ASSISTANT]: Thought: Both automated quantification and SUV calculation failed due to a coordinate system error in the DICOM metadata. To
fulfill the user's request for a report, I will generate MIP images to visually identify the lesions and then use the Vision LLM to
interpret the findings and draft the report.
Action: GENERATE_MIP(pet_uid, output_name="lung_cancer_mip")

[ASSISTANT]: Thought: Automated quantification failed due to DICOM metadata issues. I have generated MIP images and will now use the
Vision LLM to visually identify the lung lesions and draft the full quantitative report based on visual assessment.
```
